# A novel patient-level computational model of atrial fibrillation patterns and clinical outcomes for evaluating screening strategies

**DOI:** 10.1101/2024.09.30.24314669

**Authors:** Minsi Cai, Cristian Barrios-Espinosa, Michiel Rienstra, Harry J.G.M. Crijns, Ulrich Schotten, Jordi Heijman

## Abstract

**Background:** The dynamic, heterogenous nature of atrial fibrillation (AF) episodes and poor symptom-rhythm correlation make early AF detection challenging. The optimal screening strategy for early AF detection and its role in stroke prevention in different subpopulations are unknown. We developed a computational patient-level AF model able to simulate the dynamic occurrence of AF and AF-associated outcomes during the entire lifetime of a virtual patient cohort to elucidate the impact of AF screening in virtual randomized clinical trials (V-RCTs).

**Methods:** The Markov-like computer model has 7 clinical states (sinus rhythm, symptomatic/asymptomatic AF, each with/without previous stroke, and death). AF-related atrial remodeling was incorporated, which influenced the age-/sex-dependent transition probabilities between states. Model calibration/validation was performed by replicating a wide range of clinical studies. AF screening strategies and stroke rates in the presence of defined interventions were assessed in V-RCTs.

**Results:** The model simulates the entire lifetime of virtual patients with minute-level resolution and provides perfect information on the occurrence of AF episodes and clinical outcomes (stroke, death). It replicates numerous age/sex-specific episode- and population-level AF metrics, including AF incidence/prevalence, progression rates, burden, episode duration, and stroke/mortality incidence. The benefits of intermittent AF screening in V-RCTs were frequency- and duration-dependent, with systematic thrice-daily single-ECG recordings providing the highest detection rates (64.6% of AF patients being diagnosed before their symptom-based clinical diagnosis). Screening groups had comparable 5-year stroke rates and lower 25-year stroke rates than the control group. These differences were increased by more effective anticoagulation therapy, in patients with higher baseline stroke risk, or in patients with delayed clinical AF diagnosis.

**Conclusions:** We present a novel computational patient-level AF model consistent with a large body of real-world data, enabling for the first time the systematic assessment of AF-management strategies. Screening protocols with more frequent and longer monitoring have higher AF-detection rates, but stroke reduction in screening-detected individuals is highly dependent on patients’ and healthcare-systems’ characteristics. V-RCTs suggest that frequent AF screening with effective anticoagulation can reduce stroke incidence in patients with high likelihood of delayed AF diagnosis.

## Introduction

Atrial fibrillation (AF) is the most common cardiac arrhythmia with 59.7 million prevalent cases estimated worldwide in 2019.^1^ The lifetime risk of AF for individuals with European ancestry has been reported as 1 in 3 at the index age of 55 years.^2^ The prominent increase in AF is attributable to the growth of the aging population, increasing prevalence of comorbidities and better detection opportunities.^2^ AF is associated with higher risk of stroke/systemic embolism, heart failure, and mortality,^3–5^ and early detection and treatment has been shown to improve outcomes.^6^ However, early detection and treatment of AF are challenging in clinical practice, due to the highly variable duration of AF episodes, heterogenous dynamic AF progression and poor symptom-rhythm correlation.^7,8^ The optimal screening strategy to promote early AF detection and the benefits of anticoagulation therapy in patients with screening-detected AF in the general population remain a topic of debate.^9,10^ More studies are needed to optimize AF screening strategies, but such studies are challenging due to heterogeneous risk profiles in the target population, wide range of available monitoring devices, heterogenous AF patterns of patients, and high costs of clinical trials due to the low incidence of strokes.

Computational modeling potentially offers significant advantages, including perfect control over all parameters, the ability to track dynamic changes of all components, and the feasibility of comprehensively assessing various interventions. There is a long-standing history of computational modeling in cardiac electrophysiology with important real-world implications, including drug safety assessment and guidance of AF ablation through digital twins.^14,15^ However, currently available mechanistic models have so far only been used for short-term simulations (seconds). Health technology assessment models on the other hand are commonly used to assess cost-effectiveness of therapies by simulating transitions between different clinical states, each associated with different costs and values.^16,17^ While these models allow extrapolation beyond the duration of clinical trials (years) and have important implications for reimbursement decisions, these models have so far ignored AF development, pathophysiology and progression. Here, we present and validate the first computational patient-level AF model able to simulate individual AF patterns, population-level AF characteristics, key pathophysiological concepts, and relevant clinical outcomes during the entire lifetime of a virtual cohort. We employ this model to identify effective AF screening strategies and elucidate the impact of AF screening on clinical outcomes in virtual randomized clinical trials (V-RCTs).

## Methods

### Development of a dynamic patient-level Markov model

A Markov-like model was developed, comprising 7 different clinical states: sinus rhythm (SR), symptomatic and asymptomatic AF (sAF and aAF, respectively), each with or without a previous stroke, as well as death (**Figure 1A**). States were connected through age-dependent and sex-specific transition probabilities for AF development, conversion, incident stroke, and death. All virtual patients started in the SR state at birth. During each subsequent time step (30 min, unless specified otherwise), the transition probabilities toward every neighboring state were calculated. Subsequently, the state for the next time step was obtained by a stochastic process in which every state was chosen with likelihood corresponding to these probabilities **(Figure S1**). Proarrhythmic AF-related atrial remodeling was incorporated through changes in three components conceptually representing effective refractory period (ERP), atrial fibrosis, and atrial enlargement, which was determined by the time spent in AF states and age. Being in AF states decreased the value of ERP and increased the production rates of atrial fibrosis and atrial enlargement, whereas being in SR states selectively reversed the ERP variable back to normal, capturing the concept of ‘AF-begets-AF’. The relative values of the three components were used to modify the probability of AF development and cardioversion. Similarly, the non-reversible probabilities of stroke and death were modulated by vulnerability factors that increased when the virtual patient was in AF states and continued to modulate stroke/death risks after returning to sinus rhythm. A detailed model description is provided (**Supplemental Methods, Table S1, Table S2, and Figure S1**).

**Figure 1.**
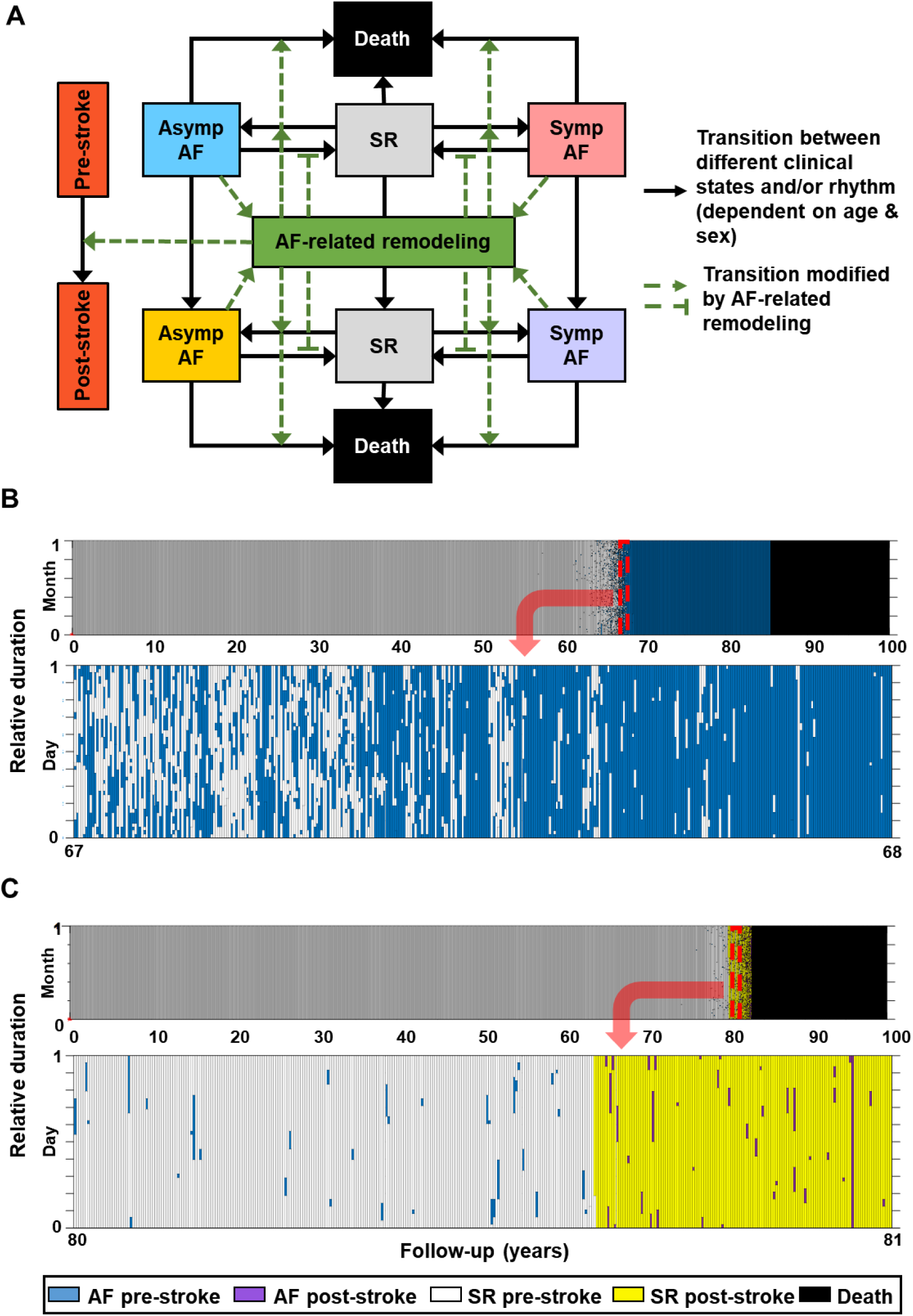
Schematic model overview and examples of AF episodes and clinical outcomes in virtual individuals simulated by the patient-level model. **A**, Patient-level model with 7 clinical states including sinus rhythm, symptomatic AF, and asymptomatic AF, each with or without previous stroke, as well as death. Transition probabilities were age/sex dependent. AF-related remodeling was also incorporated in the model as function of the time in AF, and modulated the likelihood of AF development, cardioversion, and the occurrence of stroke and death. **B**, Representative example of a patient with multiple AF episodes and AF progression during the 100-year follow-up (Top) and expanded view of the interval from 67 years to 68 years of age during which paroxysmal AF progressed to persistent AF (Bottom). **C**, Representative example of a patient with multiple AF episodes and incident stroke during the 100-year follow-up (Top) and expanded view from 80 years to 81 years of age during which the stroke event occurred (yellow-shaded area). A single column in panels **B, C** represents one month and one day in the upper and lower panels, respectively.

Inter-patient variability was simulated by setting the baseline risk of AF development and the probability of converting to sinus rhythm to a different value for each patient at the start of the simulation based on a normal distribution centered around the default parameter value, reflecting a heterogeneous genetic background in the population (**Table S1**). In addition, virtual patients were randomly assigned a male or female biological sex at the beginning of the simulation, which modulated transition probabilities throughout the simulation (**Supplemental Methods and Table S1**).

### Model calibration and validation

A general overview of the model calibration strategy is depicted in **Figure S2.** Population-level AF metrics, episode-level AF metrics, clinical outcomes, and the effects of continuous AF monitoring were calibrated by reproducing study-specific patient selection criteria (e.g., matching age/sex distribution, history/type of AF, etc., see **Table S3**) and changing parameters to reproduce the clinical outcomes. Subsequently, model validation with independent data not used for calibration was implemented by simulating a landmark analysis of stroke risks in patients with different durations of subclinical AF defined by a substudy of ASSERT (The Asymptomatic Atrial Fibrillation and Stroke Evaluation in Pacemaker Patients and the Atrial Fibrillation Reduction Atrial Pacing Trial)^18^ using the final optimized parameters.

#### Population-level metrics

Age/sex-specific AF incidence was calculated as the number of newly diagnosed AF patients divided by the number of person-years of the total population within the age interval and sex category. Age/sex-specific AF prevalence was calculated as the number of AF patients alive at the midpoint of an age interval divided by the total number of individuals alive at this moment, for men and women. The proportion of patients with paroxysmal AF (pAF) was assessed by generating a random detection timepoint. Anyone with a clinical diagnosis of paroxysmal (but not persistent) AF before this timepoint would count towards the proportion of pAF. These population-level metrics were calculated based on a virtual diagnosis of clinical AF, consistent with the epidemiological origin of the clinical data. A virtual patient received a clinical AF diagnosis in our model whenever a symptomatic AF episode reached a duration of 3 hours for the first time, reflecting a lower bound on the time needed to receive medical care. The impact of this healthcare system-related parameter was subsequently investigated in a sensitivity analysis (discussed below). The proportion of symptomatic AF patients was calculated as the number of patients with any symptomatic episodes divided by the total number of AF patients.

#### Episode-level metrics and progression

The average number of AF episodes per day (AF frequency), the maximum duration among AF episodes (maximum duration), and the percentage of the time in AF (AF burden) were calculated.^8^ Low/intermediate/high burden groups were defined as AF burden <0.5%, 0.5%-2.5%, and ≥2.5% based on the RACE-V study (Interaction Between HyperCoagulability, Electrical Remodelling, and Vascular Destabilisation in the Progression of Atrial Fibrillation registry).^19^ Clinical AF progression rate was defined as the yearly percentage of patients developing persistent or permanent AF. while AF burden progression rate was defined as the yearly percentage of patients with an increase in AF burden >3%, based on the RACE-V study.^20^ AF recurrence rate after spontaneous cardioversion was assessed using Kaplan-Meier (K-M) curves.^21^ All metrics were calibrated in virtual patients with a clinical diagnosis of pAF selected to match the characteristics of the clinical studies. The proportion of symptomatic AF episodes was calculated among all AF episodes of all virtual AF patients and compared to clinical data from patients with CRM.

#### Clinical outcome metrics

Age/sex-specific all-cause mortality was calculated as the number of new deaths divided by the number of person-years of the total population during the age range in men and women. The percentage of deaths was calculated as the number of deaths within the age interval divided by the total number of deaths. Similar definitions applied for age/sex-specific stroke incidence and percentage of strokes. Mortality and stroke rates for specific time frames were calculated as the total number of deaths and strokes during the follow-up period divided by the total population at the start of follow-up. Cumulative death and stroke rates were estimated by K-M curves.

### Parameter optimization

Fifty-eight parameters were included in the current model (**Table S1**, including ones for the sex of the virtual patient and screening characteristics such as frequency and duration of screening). Initial values were manually chosen based on the clinical data for the aforementioned metrics and a sensitivity analysis was implemented for 47 AF- and outcome-relevant parameters in order to show their impact on model calibration (**Supplemental Methods and Figure S3**). Subsequently, 17 parameters were selected for further optimization using the Nelder-Mead Simplex algorithm in Matlab R2024a (Mathworks, Natick, MA) using the difference between the metrics calculated based on model output and the clinical data.

### AF detection rate and incident strokes with continuous monitoring

The optimized model was verified by simulating the LOOP study (Atrial Fibrillation Detected by Continuous ECG Monitoring Using Implantable Loop Recorder to Prevent Stroke in High-risk Individuals).^9^ Based on the inclusion criteria in the LOOP study, several parameters were changed to simulate the 3.25-year continuous screening (the average duration of screening) in individuals >70 years with at least one additional stroke/AF risk factor but without previous stroke and clinical AF, and the effect of anticoagulation therapy that could decrease the stroke probability by 70% (**Table S4**).^22,23^ Next, AF detection rates for specific time frames with or without CRM were calculated as the total number of patients with AF episodes detected (with CRM) or newly diagnosed clinical AF (for patients with/without CRM) during the monitoring time, divided by the total population at the start of follow-up. Cumulative AF detection rates were estimated by K-M curves.

### Assessment of different AF screening strategies and their impact on stroke

Systematic screening protocols with different frequency and duration were designed and were applied to all virtual individuals from 65 years old to death, while symptom-based strategies were initiated after the first symptomatic AF episode after 65 years of age (independent of its duration) for 2 weeks (short-term symptom-based strategies) or until death (long-term symptom-based strategies) (**Table S5**). Whenever clinical AF was diagnosed before or during follow-up, screening protocols for the virtual individual were stopped. Screening-detected AF was defined as patients without clinical AF but with at least one AF episode detected by the screening. Earlier-detected AF was defined as patients with screening-detected AF who also met the criteria for a subsequent clinical AF diagnosis. In all analyses, virtual anticoagulation therapy was initiated as soon as AF was detected by implementing a 70% reduction in the probability of stroke (i.e., parameter[51] set to 0.3). Subsequently, 5- or 25-year stroke rates after the initiation of AF screening were compared among different strategies. The effect of screening was assessed for different stroke risks (controlled by parameter[17]), defined as baseline stroke risk and 3 times baseline stroke risk. Additional sensitivity analyses were conducted by changing the cutoff value for a clinical AF diagnosis from 3 hours to 24 hours of continuous symptomatic AF, and by increasing the efficacy of anticoagulation therapy from 70% to 90% less stroke incidence, reflecting potential future advances in anticoagulation therapy.

### Statistical description of metrics

Five independent datasets were simulated using the optimized parameters to produce calibration bar plots with error bars (mean ± standard deviation) for all metrics. For K-M curves, all datasets were combined together and plotted with 95% confidence intervals (95% CIs) calculated by Greenwood’s method. Hazard ratios (HRs) with 95%CIs in model validation were estimated by Cox regression. Mean relative reduction of stroke for each AF screening strategy compared with the control group was estimated with 95% CIs using 10 simulated datasets due to the limited number of strokes at 5-year follow-up. All K-M plots and survival analysis were conducted in RStudio v2023.06.1 and statistical software R v4.3.0 (R core team, 2023)

## Results

### Population-level AF metrics

The patient-level model was able to simulate the entire lifetime of a virtual cohort of 10,000 individuals (50% female). During the 100-year follow-up, the virtual patients showed various AF patterns and clinical outcomes including death and stroke (top panels in **Figure 1B and 1C**). All AF episodes were captured with 30-minute-level resolution (bottom panels in **Figure 1B and 1C**), and in a subset of individuals showed the expected dynamic progression from pAF to persistent AF (**Figure 1B**).

Consistent with the known age dependence of AF risk, the AF prevalence (**Figure 2A**) and incidence (**Figure 2B**) in this virtual cohort were significantly higher in older age groups. Although AF incidence was numerically consistent with the range estimated from numerous studies,^24–37^ AF prevalence was slightly overestimated in the virtual cohort, possibly due to underestimation of clinical AF in the real-world general population or the definition of clinical AF in the model. Sex-specific AF prevalence and incidence were also calibrated and showed a similar age-dependence, with both metrics significantly higher in males, especially in the older age groups (**Figure S4**), consistent with clinical data.^27–31,33,35–38^ The proportions of pAF (vs. non-pAF) patients (**Figure 2C**) and symptomatic (vs. asymptomatic) AF patients (**Figure 2D**) were also consistent with clinical data^39,40^ and accounted for around 50% and 75% of total AF patients, respectively.

**Figure 2.**
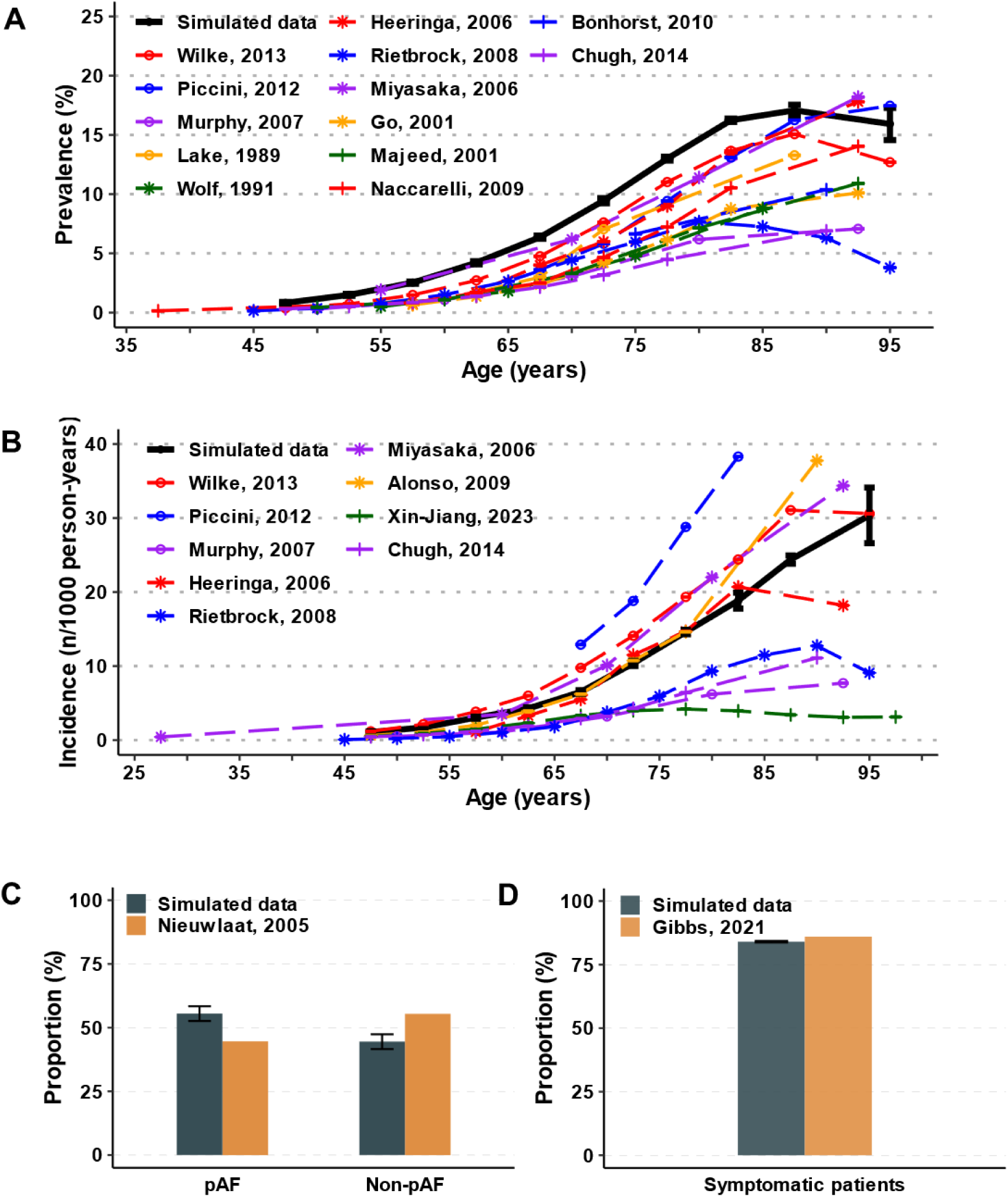
Comparison of population-level atrial fibrillation (AF) metrics in 10,000 virtual individuals to epidemiological data. **A-B**, Age dependence of the prevalence (A) and incidence (B) of clinical AF in model (black line) and different clinical studies (colored lines).^24–36^ Sex-specific AF prevalence and incidence are shown in **Figure S4**. **C**, The proportion of paroxysmal AF (pAF) and non-paroxysmal AF in virtual patients with a clinical diagnosis of AF compared to clinical data from Nieuwlaat et al.^40^ **D**, The proportion of symptomatic AF patients in the model compared to Gibbs et al.^39^

### AF-episode and progression metrics

In pAF patients, the AF frequency, maximum duration of AF episodes, and AF burden during a 2-week period were assessed. Although the model slightly underestimates AF frequency (**Figure 3A**), likely due to the temporal resolution (30 minutes) used in current simulations, the maximum duration of AF episodes (**Figure 3B**) and AF burden (**Figure 3C**) were in good agreement with the real data.^8^ Consistent results were also obtained specifically for male and female patients (**Figure S5**).

**Figure 3.**
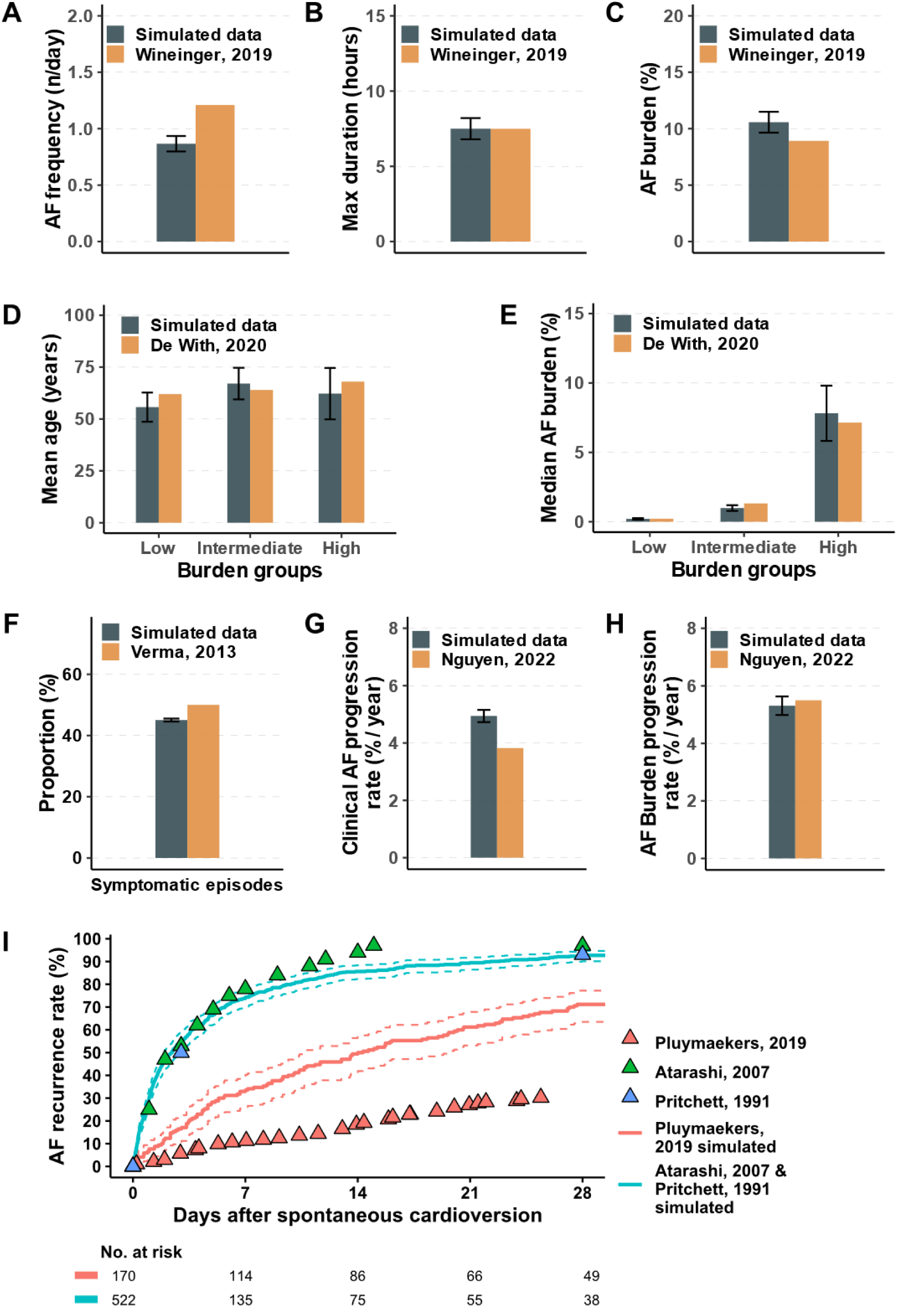
Comparison of episode-level atrial fibrillation (AF) metrics and AF progression in the patient-level model to rhythm-monitoring data in AF patients. **A-C**, Frequency (**A**), maximum episode duration (**B**) and AF burden (**C**) during a 12-day follow-up period in patients with paroxysmal AF compared to Wineinger et al.^8^ D-E, Mean age (**D**) and median AF burden (**E**) in different AF burden groups defined by the RACE-V study.^19^ F, The proportion of symptomatic AF episodes from all virtual patients with AF compared to implantable cardiac monitor data.^41^ G-H, Yearly progression rate from paroxysmal to non-paroxysmal AF (**G**) or AF burden progression rate (**H**) compared to data from the RACE-V study.^20^ **I**, AF recurrence rate after spontaneous cardioversion in patients with early phase AF, as defined by the different clinical trials.^21,43,44^

We subsequently reproduced the selection criteria for early pAF from the RACE-V study.^19^ The resulting mean age in patients with low, intermediate, and high AF burden according to the RACE-V definition was broadly consistent with the clinical data, ranging from 62 to 68 years (**Figure 3D**), and simulated median AF burden was 0.2%, 1.0%, and 7.8% respectively (**Figure 3E**), comparable to 0.2%, 1.3% and 7.1% observed in the clinical study.^19^ The proportion of symptomatic AF episodes was 45%, similar to the observed data (50%) in patients with continuous monitoring (**Figure 3F**).^41^

AF is known as a progressive disease. While the clinical AF progression rate from pAF to persistent AF was slightly overestimated at ∼4.9% / year versus 3.8% / year (**Figure 3G**),^20^ this factor can be quite variable depending on clinical characteristics and AF detection parameters.^42^ AF burden progression rate is a more robust parameter and agreed well between simulated data (5.3% / year) and clinical data (5.5% / year; **Figure 3H**).^20^

AF recurrence after spontaneous cardioversion varies significantly between pAF patients with different baseline characteristics, ranging from 30%-90%.^21,43,44^ Therefore, the AF recurrence rate in the virtual cohort was calibrated by reproducing baseline characteristics (age, history of AF) in individual simulations for each clinical dataset. Different pattern of recurrent AF episodes during the 28-day follow-up were documented for each virtual patient, with sparse and short episodes in patients with low AF recurrence rate (**Figure S6A**), but frequent and relatively longer episodes in patients with higher AF recurrence rate (**Figure S6B**). During the 28-day follow-up, recurrent AF episodes were detected in 60-90% of virtual patients with pAF (**Figure 3I**), which was in agreement with clinical data from some, but not all, randomized trials.^21,43,44^ The difference between simulated recurrence and clinical data might in part be explained by the declining patients’ adherence to a full monitoring during the follow-up.^45^ Together, these data confirm that the model can reproduce a wide range of AF characteristics occurring during the entire lifetime of a virtual population.

### Clinical outcomes

AF is associated with increased morbidity and mortality, notably due to an increased risk of heart failure and stroke.^2–5^ The ultimate goal of AF management is to improve clinical outcomes. Accordingly, we assessed clinical outcomes in individuals with or without AF in our model. Mortality (**Figure 4A**), the percentage of deaths per age category (**Figure 4B**), stroke incidence (**Figure 4C**), and the percentage of strokes per age category (**Figure 4D**) in the virtual cohort were all consistent with age-dependent data from literature.^46–48^ A similar age dependence was present for both females and males, with higher risks of death (**Figure S7A-D**) and stroke (**Figure S8A-D**) in males, consistent with clinical data. Furthermore, the acute, out-of-hospital death (≤24 hours) and early-phase fatality (≤30 days and within 31-365 days) after stroke were reproduced for the total population (**Figure S9A**), as well as in female (**Figure S9B**) and male (**Figure S9C**) subpopulations.^49,50^

**Figure 4.**
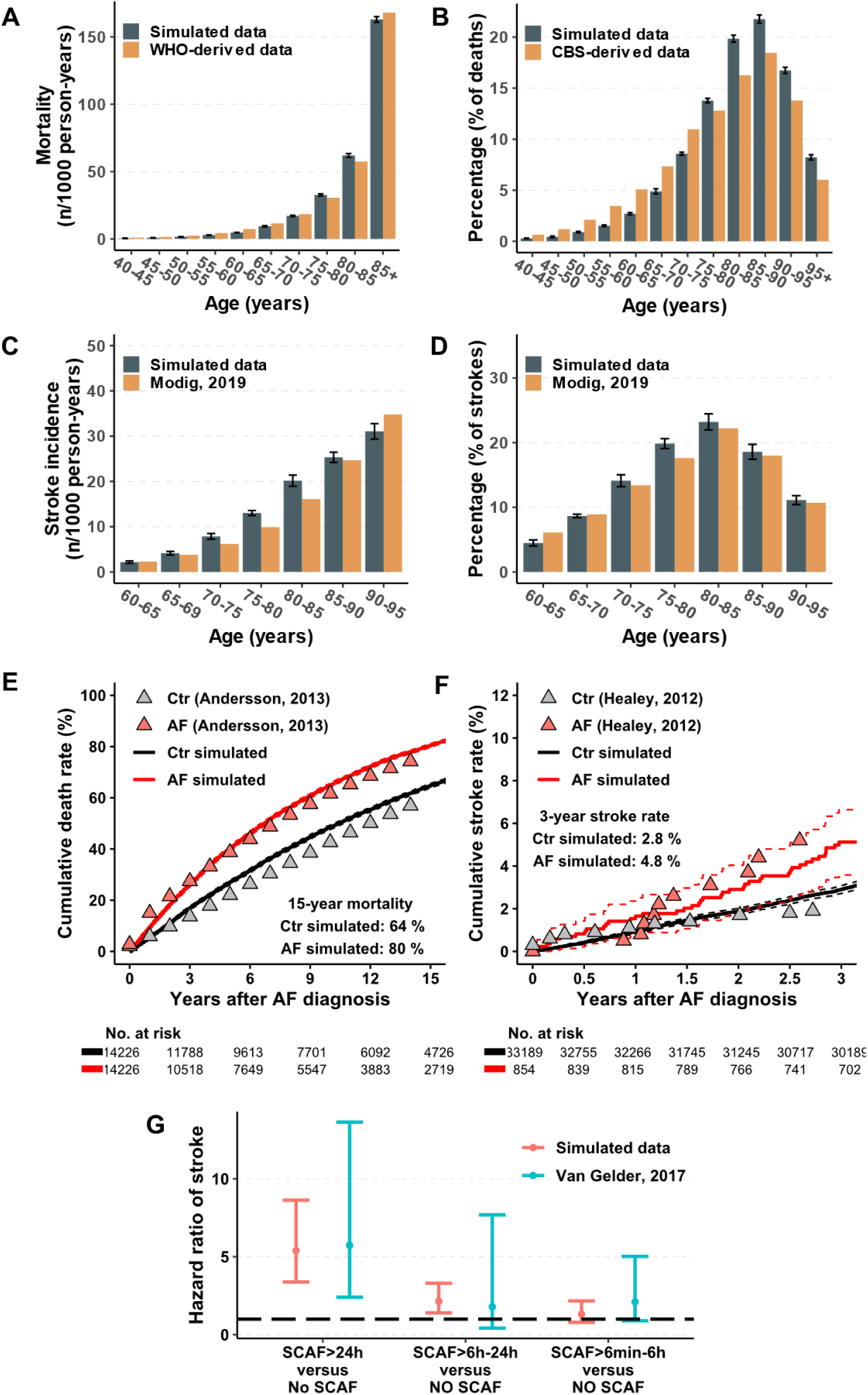
Comparison of clinical outcomes in 10,000 virtual patients to epidemiological data. **A**, Mortality across different age groups in the virtual cohort compared to the data from WHO.^47^ **B**, Percentage of deaths across different age groups in the virtual cohort compared to the data from Statistics Netherlands.^48^ **C-D**, Stroke incidence (**C**) and percentage of strokes (**D**) across different age groups in the virtual cohort compared to the data from Modig et al.^46^ **E**, Mortality in patients with clinical AF compared to age- and sex-matched patients in model and data from Andersson et al.^51^ **F**, Cumulative stroke rate in patients with or without any AF episodes compared to the continuous monitoring data from ASSERT study.^52^ **G**, Validation of stroke risks in patients with different durations of subclinical AF compared to patients with no subclinical AF in the model compared to data from the ASSERT study,^18^ which were not used for model development and calibration.

Age- and sex-matched mortality at 15 years after AF diagnosis was approximately 25% higher in simulated AF patients than in simulated individuals with SR (**Figure 4E**), which aligned well with clinical data.^51^ Similarly, an almost 2-fold higher risk of cumulative incident strokes after AF diagnosis was observed in both simulated AF and clinical AF cohorts compared with those in SR over a 3-year follow-up period (**Figure 4F**).^52^

After calibrating the model to accurately reproduce these age- and sex-dependent clinical outcomes, we validated the model by comparing stroke risks in patients with different durations of AF episodes to a clinical dataset that was not used for parameter estimation. The simulated stroke risks in patients with AF over a 3.5-year follow-up resulted in HRs of 5.40 (95%CI: 3.38-8.63), 2.15 (95%CI: 1.40-3.30) and 1.31 (95%CI: 0.79-2.17) compared to individuals without AF for episodes of >24 hours, 6-24h and 6 minutes-6 hours, respectively. These numbers were highly consistent with 5.73 (95%CI: 2.41-13.64), 1.79 (95%CI: 0.42-7.69) and 1.31 (95%CI: 0.89-5.02) observed in the ASSERT study (**Figure 4G**).^18^ These validation results highlight the model’s accuracy and predictive abilities.

### Impact of different screening strategies on AF detection and stroke reduction

AF screening has the potential to enable early AF detection and initiation of anticoagulation therapy to reduce stroke risk, but the optimal screening strategy and the benefit of anticoagulation for screening-detected AF remain uncertain.^9,10^ Firstly, the model’s suitability for evaluating AF screening was assessed by simulating the detection rates and benefits for stroke prevention of CRM based on the LOOP study.^9^ Cumulative stroke rates (**Figure 5A**) and AF detection rates (**Figure 5B**) reproduced in virtual CRM and control groups were highly similar to the clinical data, with 5.5-year stroke rates of 4.1% and 5%, and 5.5-year AF detection rates of 29.5% and 16.5% in virtual CRM and control groups, respectively. This confirmed that the model can be used to assess the impact of screening on AF detection and stroke reduction.

**Figure 5.**
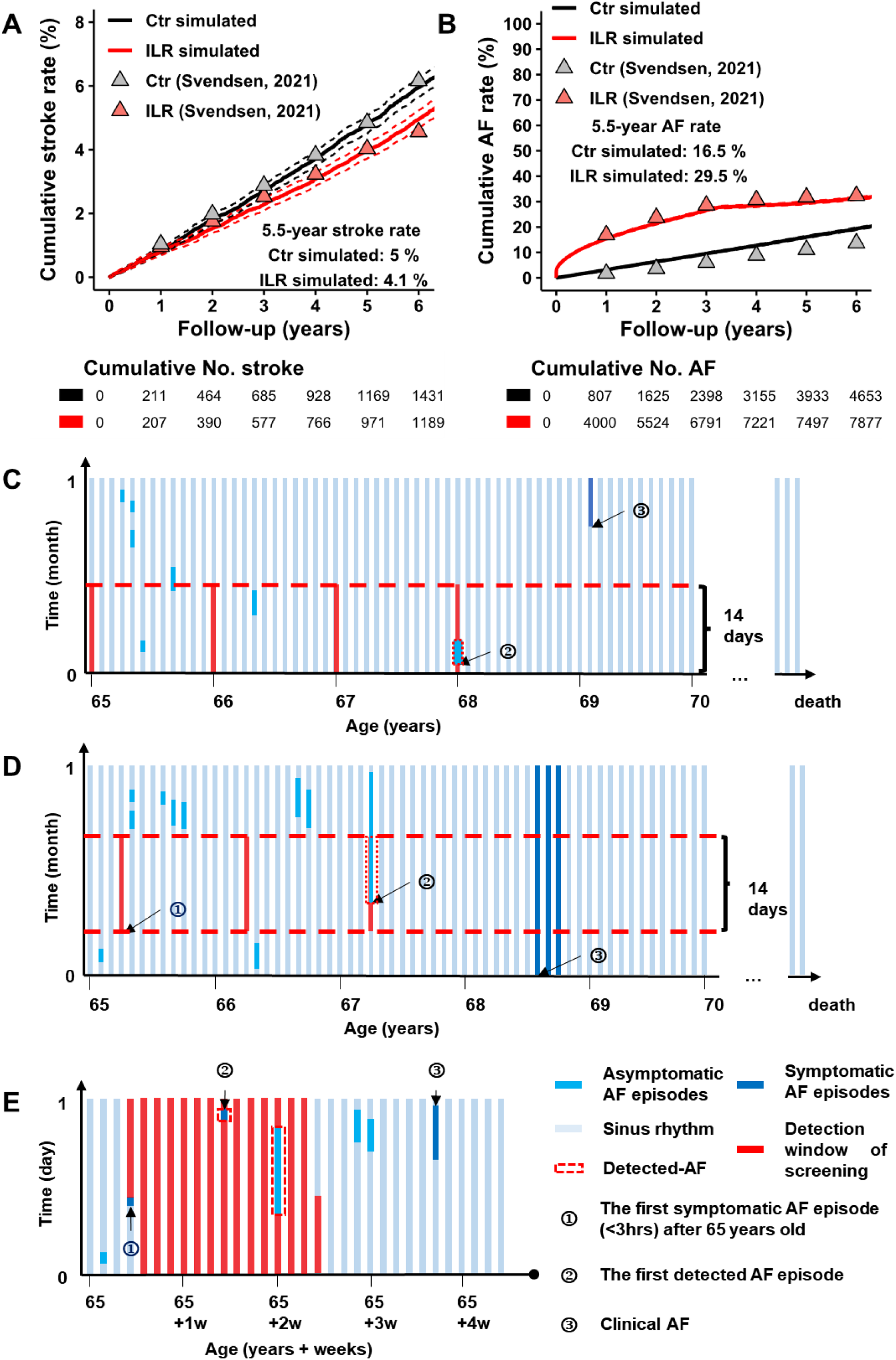
AF screening in 10,000 virtual patients with or without continuous rhythm monitoring (CRM) and schematic illustration of self-defined AF screening protocols. **A-B**, Cumulative stroke rates (**A**) and AF detection rates (**B**) with or without CRM in the virtual cohort compared to data from patients with implanted loop recorders from the LOOP trial.^9^ **C-E**, Schematic illustration of self-defined systematic AF screening (**C**), symptom-based long-term (until death, **D**) or short-term (2 weeks, **E**) AF screening.

Because the follow-up time of CRM is limited by battery life and it will not be feasible to employ CRM for all patients at risk of AF, we employed the model to evaluate self-defined systematic (**Figure 5C**) and symptom-based AF screening protocols (**Figure 5D and 5E**). Generally, CRM and intermittent strategies with higher frequency and longer monitoring periods identified more AF cases from the total population (**Table 1**). The top 5 screening protocols that patients with AF would benefit from (defined as percentage of AF patients with screening-detected AF before a clinical diagnosis) were systematic CRM (99.7%, 3524 / 3534), systematic daily 3-time single ECG (64.6%, 2268 / 3511, representing a smart-watch-based approach), long-term (until death) symptom-based CRM (52.5%, 1852 / 3525), long-term symptom-based daily 3-time single ECG (35.1%, 1217/3464), and systematic yearly 14-day Holter monitoring (23.3%, 818/3517). AF could be detected earlier than the clinical diagnosis by 720 (IQR: 140, 1448), 301 (IQR: 9, 697), 390 (IQR: 94, 813), 204 (IQR: 11, 551), and 285 (IQR: 66, 718) days, respectively, in these protocols (**Table 1**). Finally, we assessed the impact of the 5 screening strategies on stroke rates in V-RCTs, comparing oral anticoagulation upon routine clinical AF diagnosis (control group) with oral anticoagulation upon screening-detected or clinical AF. At 5-year follow-up, all strategies showed similar stroke rates to the control group (**Figure 6A**). However, at 25-year follow-up stroke rates were slightly reduced with the lowest stroke rate (5% relative risk reduction) with systematic CRM (**Figure 6B**).

**Figure 6.**
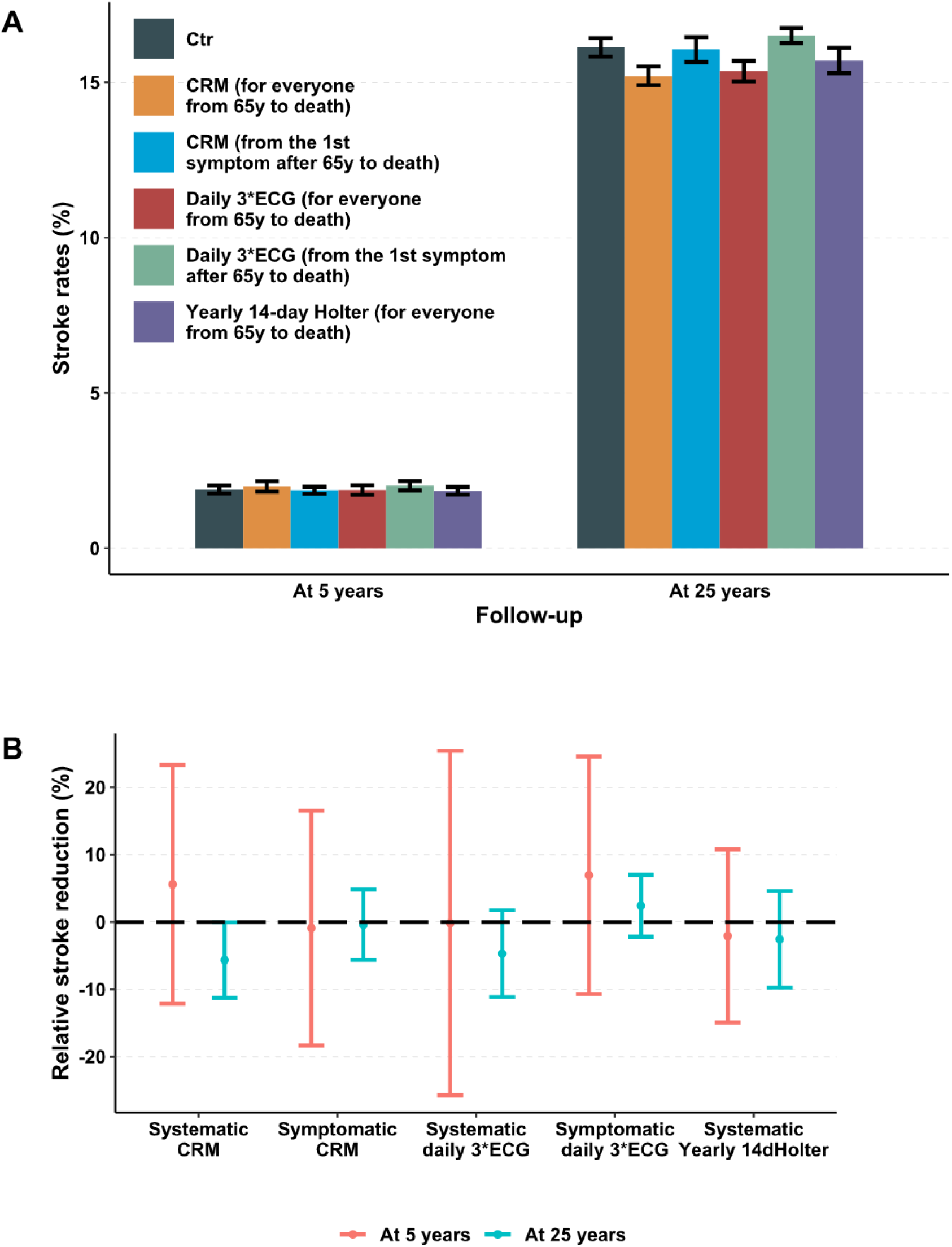
Stroke rates in populations with or without AF screening. **A**, Comparison of 5-year and 25-year stroke rates in populations with normal baseline stroke risk and different AF screening strategies. **B**, The 5-year and 25-year relative stroke reduction for AF screening groups with normal baseline stroke risk and anticoagulation therapy with 70% efficacy, compared with the control group. Here, systematic screening means AF screening initiated for everyone from 65 years to death, and symptomatic screening means AF screening initiated from the 1^st^ symptomatic AF episode after 65 years to death. The diagnostic criterion for clinical AF was 3 hours of symptomatic AF. See **Methods** for details.

**Table 1.** AF detection rates of different screening protocols applied in the virtual cohort

| Protocols applied (10,000 individuals) | % of patients benefitting from screening (n/ AF cases possibly at benefit*) | % of screening-detected AF<sup>†</sup> (n/ AF cases possibly at benefit) | % of earlier – detected AF<sup>‡</sup> (n/ AF cases possibly at benefit) | # of days ahead of clinical AF diagnosis<sup>§</sup> for earlier-detected AF (n, IQR) |
| --- | --- | --- | --- | --- |
| **Continuous AF screening (systematic or long-term symptom-based)** |  |  |  |  |
| Systematic continuous screen | 99.7%<br>(3524 / 3534) | 31.6%<br>(1118 / 3534) | 68.1%<br>(2406 / 3534) | 720 (140, 1448) |
| Symptom-based continuous screen | 52.5%<br>(1852 / 3525) | 8.0%<br>(281 / 3525) | 44.6%<br>(1571 / 3525) | 390 (94, 813) |
| **Systematic AF screening (all participants from 65y to death)** |  |  |  |  |
| Yearly 1min ECG | 4.7%<br>(165 / 3491) | 4.7%<br>(164 / 3491) | 0.03%<br>(1 / 3491) | 516 |
| Daily 3 * 1min ECG | 64.6%<br>(2268 / 3511) | 17.2%<br>(603 / 3511) | 47.4%<br>(1665 / 3511) | 301 (9, 697) |
| Yearly 7d Holter | 16.7%<br>(588 / 3514) | 8.6%<br>(301 / 3514) | 8.2%<br>(287 / 3514) | 248 (53, 605) |
| Yearly 14d Holter | 23.3%<br>(818 / 3517) | 10.4%<br>(365 / 3517) | 12.9%<br>(453 / 3517) | 285 (66, 718) |
| **Short-term symptom-based AF screening (from the 1<sup>st</sup> symptomatic AF after 65y, for 2w)** |  |  |  |  |
| Daily 1min ECG | 0.2%<br>(8 / 3539) | 0%<br>(0 / 3539) | 0.2%<br>(8 / 3539) | 428 (163, 558) |
| Daily 3 * 1min ECG | 0.5%<br>(26 / 3517) | 0%<br>(0 / 3517) | 0.5%<br>(26 / 3517) | 100 (36, 277) |
| Weekly 1d Holter | 0.6%<br>(20 / 3400) | 0.06%<br>(2 / 3400) | 0.5%<br>(18 / 3400) | 218 (107, 647) |
| One-time 14d Holter | 2.7%<br>(97 / 3583) | 0.4%<br>(16 / 3583) | 2.3%<br>(81 / 3583) | 279 (111, 610) |
| **Long-term symptom-based AF screening (from the 1<sup>st</sup> symptomatic AF after 65y, until death)** |  |  |  |  |
| Daily 1min ECG | 17.6%<br>(623 / 3530) | 1.6%<br>(58 / 3530) | 16.0%<br>(565 / 3530) | 183 (17, 410) |
| Daily 3 * 1min ECG | 35.1%<br>(1217 / 3464) | 4.0%<br>(139 / 3464) | 31.1%<br>(1078 / 3464) | 204 (11, 551) |
| Yearly 1d Holter | 1.2%<br>(29 / 3558) | 0.2%<br>(1 / 3558) | 1.1%<br>(28 / 3558) | 167 (46, 383) |
| Yearly 14d<br>Holter | 12.0%<br>(429 / 3563) | 1.2%<br>(41 / 3563) | 10.9%<br>(388 / 3563) | 229 (37, 507) |

Results are presented as absolute numbers with percentage or median numbers with quartiles.
\* AF cases possibly at benefit are those with **ANY** AF episodes during **LIFETIME** follow-up and still alive at 65 years of age (i.e., start of screening) without clinical AF diagnosis (= the number of total AF cases within 100 years - AF cases died before 65y - clinical AF cases before 65y)
† Screening-detected AF are patients detected by screening protocols **ONLY**
‡ Earlier-detected AF are patients detected by screening protocols earlier than their clinical AF diagnosis
§ Clinical AF was diagnosed when the first symptomatic AF episode lasted $\geq 3$ hours

Given the limited benefits from AF screening in an unselected population, we finally performed a sensitivity analysis to assess the influence of the moment of clinical AF diagnosis, the baseline stroke risk, and the efficacy of anticoagulation therapy (**Figure 7**). The 5-year stroke rates were similar in all groups and were not significantly reduced compared with the control group (all 95% CIs cross the zero-effect line). However, different AF screening strategies showed distinct relative stroke risk reduction at 25-year follow-up, the extent of which was sensitive to the efficacy of anticoagulation therapy, baseline stroke risk, and the latency for a clinical AF diagnosis (**Figure 7 and Figure S10-12**). The largest effects were observed with systematic CRM, daily 3 times single ECG, and yearly 14-day Holter groups with significant 6-12% reductions in stroke incidence (**Figure 7**).

**Figure 7.**
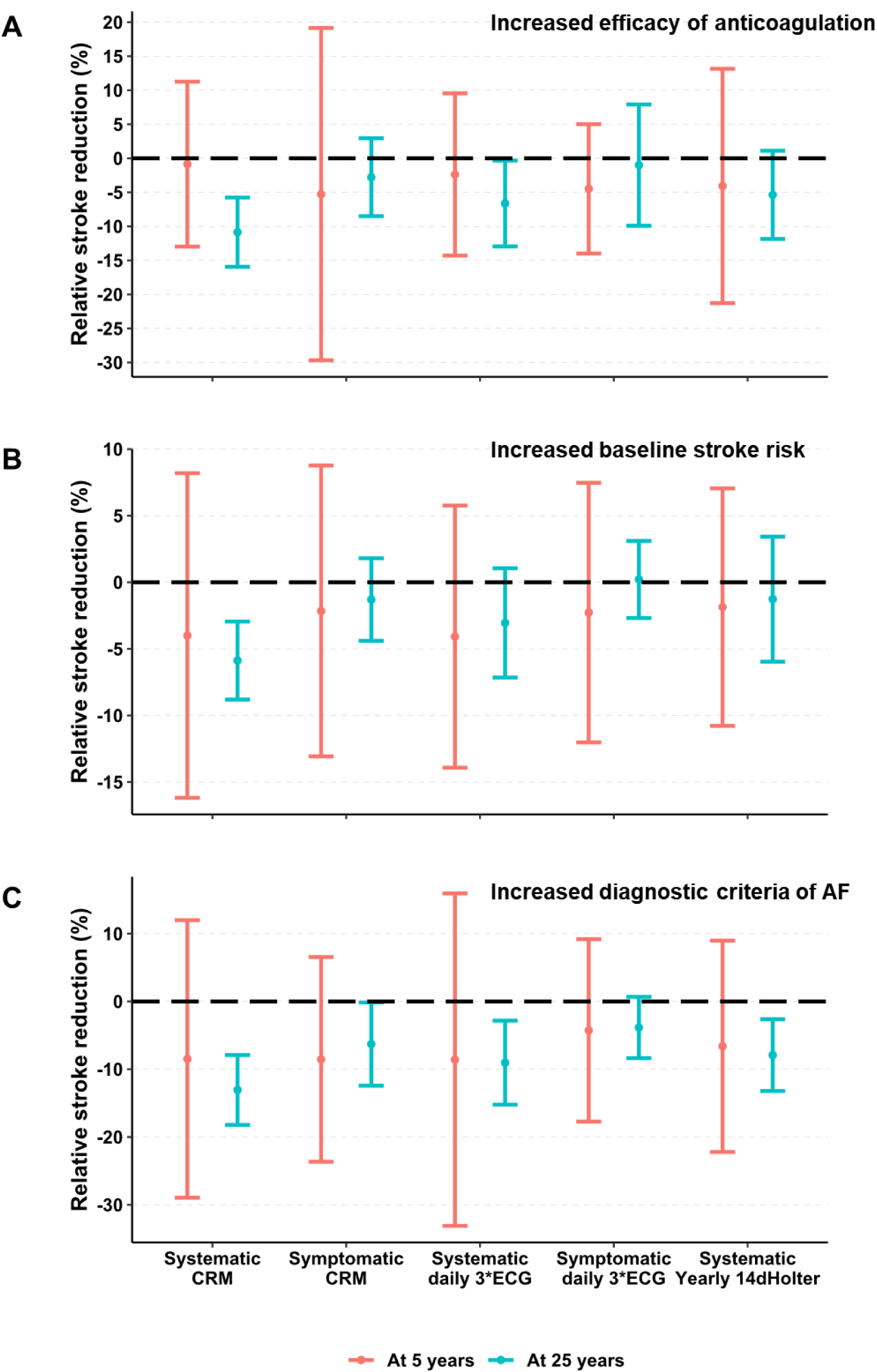
Sensitivity analysis of the impact of different healthcare factors on relative stroke risk reduction induced by AF screening. **A-C**, The 5-year and 25-year relative stroke reduction for AF screening groups with increased efficacy of anticoagulation therapy (90% less stroke compared with the control group (**A**), 3-fold increase in baseline stroke risk (**B**), or with a diagnostic criterion of clinical AF of 24 hours symptomatic AF (**C**). Abbreviations of different screening strategies are the same as in Figure 6.

## Discussion

We presented a novel patient-level AF model that can simulate the entire lifetime of a virtual individual, capturing individual AF episodes, AF progression and relevant clinical outcomes with minute-level resolution. The model incorporates key pathophysiological components, including ‘AF begets AF’ and highlights the complex, heterogeneous and dynamic development and progression of AF. It reproduces many important population-level and episode-level AF characteristics, including their age- and sex dependence. The model enables for the first time a systematic evaluation of AF screening strategies and their impact on clinical outcomes in virtual RCTs, showing huge heterogeneity in AF detection rates and identifying determinants of stroke reduction when anticoagulation therapy after clinical or screening-detected AF was simulated, such as the efficacy of anticoagulation therapy, the baseline stroke risk profile, and the latency of clinical AF diagnoses.

### Computational models of atrial fibrillation

Mechanistic modeling, data-driven modeling and health-technology assessment modeling are traditionally different approaches for computational modeling of AF, each with a long history and several important real-world applications, including the guidance of AF ablation using digital twins, the prediction of a history of AF from a sinus rhythm ECG and the assessment of cost-effectiveness of novel therapies.^14^ However, none of these approaches have simulated AF development, progression and its associated outcomes, making them unsuitable for the benefit-assessment of AF screening. Chang et al. were the first to develop a patient-level Markov model for the simulation of AF progression during a person’s lifetime.^53^ However, this model did not include clinical outcomes and AF incidence, characteristics and progression were not calibrated based on real-world data. Another study employed a patient-level Markov model to assess the effectiveness of 45 screening strategies by simulating bleeding and stroke risks in the general population. AF characteristics, common comorbidities, anticoagulation therapy, and screening specificity were considered. However, no individual AF episodes were simulated, and the progressive nature due to AF-induced remodeling was ignored in these simulations.^54^ To the best of our knowledge, our model is the first to incorporate the information about every single symptomatic or asymptomatic episode, together with electrical and structural remodeling of the atrium and clinical outcomes during the entire lifetime with minute-level resolution, providing all the components required for the systematic assessment of AF screening strategies.

### Determining optimal AF screening strategies

The LOOP trial showed that CRM results in a >2-fold increase in AF detection.^9^ However, AF screening with implantable CRM devices is not feasible in the general population. After successful replication of the results from LOOP study, we therefore evaluated various self-designed AF screening protocols. Our results suggest that systematic screening (assessing the whole population from a certain age onwards) outperforms symptom-based screening (only starting screening in individuals with suspected symptomatic AF episodes). This can be explained by the fact that 1 in 4 AF patients are totally asymptomatic and asymptomatic AF episodes account for 50% of total episodes in the virtual cohort, in line with clinical data.^39,41^ On the other hand, the AF detection rates of both systematic and symptom-based AF screening strategies are frequency- and duration-dependent in the simulation, which is consistent with previous simulation data^55^ and suggests a need for long-term monitoring for better AF diagnosis. This idea is supported by a recent RCT that one-time single-lead ECG screening at 65 years or older does not increase the rate of new AF diagnosis significantly in primary-care settings.^56^ Of note, longer total screening time is not associated with higher AF detection rates. For example, systematic daily 3-times 1-min ECG (1095 minutes per year) detects more AF patients than a systematic yearly 14-day Holter (20160 minutes per year). This result is likely due to the typical patterns of AF development, often beginning with sporadic and short-lasting AF episodes and then progressing to more frequent and longer AF episodes during the follow-up (**Figure 1B and 1C**). In the current simulation, AF screening will be initiated at 65 years, when most virtual patients are most likely in the early phase of AF development. Thus, different screening strategies may capture this dynamic AF progression with heterogenous sensitivity. Our results indicate that heterogenous AF development and progression in the target population should be considered when choosing the best AF screening strategy. Long-term short, frequent monitoring facilitated by developments in single-lead ECG hardware, e.g., in smart-watch devices, may be more sensitive to identify sparse AF episodes. The feasibility of AF screening with consumer devices has been validated in the general population, showing improved anticoagulation therapy, rate- or rhythm-control strategies and risk factor management in diagnosed AF patients.^57^

### Potential benefits of AF screening and anticoagulation therapy

After the evaluation of AF detection rates of different screening strategies, we evaluated the potential benefits of virtual anticoagulation therapy for screening-detected AF. The 5-year stroke rates are similar between control and screening groups, and only systematic CRM shows slight relative stroke risk reduction during long-term (25-year) follow-up in the unselected baseline population over 65 years of age. These results are consistent with recent RCTs which have shown small net reduction or no reduction in stroke despite increased AF diagnosis in individuals with extra stroke risks but without previous AF diagnosis undergoing intermittent screening or CRM during 5-7 years follow-up.^9,10^ These results support the idea that although AF screening will identify more AF cases and lead to earlier anticoagulation therapy, benefits of AF screening in terms of stroke reduction may still be limited, even when long-term CRM is used. Considering the significantly higher risk of major bleeding in patients with device-detected subclinical AF and anticoagulation therapy^13^ and the potential costs of such a screening strategy, screening for everyone is likely not the best choice. We therefore employed the model to evaluate parameters that may affect the benefit of AF screening.

In our sensitivity analysis, patients with more effective anticoagulation therapy, higher baseline stroke risk, and higher likelihood of delayed clinical AF diagnosis, e.g., due to poor ad-hoc accessibility to cardiology care (simulated by prolonged AF-duration criteria for clinical AF diagnosis), the reduction of 25-year stroke rates was more significant, particularly in CRM and systematic screening groups **(Figure 6, Figure 7, and Figure S10-S12**). Thus, our approach suggests that tailored systematic screening approaches in selected settings could be beneficial. One recent population-level simulation has indicated that AF screening can result in quality-adjusted life-year gains and a reduction in stroke risks, which varies significantly and is highly dependent on specific combinations of strategies used.^54^ Of note, most current AF-screening studies, including our simulations, have only evaluated the screening-enabled benefits of anticoagulation therapy, whereas a systematic evaluation of the combined effects with other interventions such as rhythm control and upstream treatments has not been performed.

### Potential limitations

Although age, the most important risk factor for AF development, and sex have been explicitly included in the model, other known AF risk factors such as hypertension and heart failure are only incorporated implicitly by reproducing epidemiological data sets. In particular, the model captures the age dependence of AF-development and mortality. Virtual individuals that develop AF or die at a young age likely implicitly represent those people with a higher burden of risk factors and comorbidities, even when these are not modelled explicitly. Nevertheless, the absence of these components can make the exact replication of inclusion criteria in different studies challenging. We have tried to simulate the study-specific patient selection as much as possible based on age, sex, AF history and clinical priors (**Table S3**). While the model is generally consistent with a wide range of clinical data, some quantitative differences exist and may be due to this patient selection. On the other hand, results from clinical studies may also be biased due to various practical issues during the implementation. For example, a lack of patients’ adherence to full monitoring during the 4-week follow-up after spontaneous cardioversion was reported,^45^ but without sufficient detail to reproduce in the model, potentially contributing to the mismatch between simulated and observed AF recurrence rates (red line and triangles in **Figure 3I**). In addition, the model uses a temporal resolution of 30 minutes for computational efficiency. As such, short episodes of micro-AF cannot be simulated, potentially contributing to the underestimation of the AF frequency (**Figure 2A**). However, the clinical relevance of such short-lived episodes remains uncertain at present. Finally, only stroke and death were incorporated as clinical outcomes in our model. Thus, safety analyses, e.g., in terms of bleeding risk after starting anticoagulation in screening-detected AF,^13^ and risk-benefit assessment cannot be simulated in the current V-RCTs and should be considered in future research. Nevertheless, our simulation results are consistent with the limited short-term efficacy of oral anticoagulation therapy for screening-detected AF reported in recent trials. They also help to identify conditions when AF screening may benefit long-term outcomes, which will ultimately need to be validated in a real-world RCT.

### Future perspectives

The current computational patient-level AF model provides perfect control over every component responsible for AF development, progression and relevant clinical outcomes and can reproduce numerous population-level metrics and essential clinical characteristics of AF during the entire lifetime of a virtual patient. As such, the model is expected to be helpful for future mechanistic studies on AF development, evaluating AF pattern changes and progression, predicting long-term clinical prognosis, and assessing the efficacy of different treatments in different populations. As such, the model may provide valuable information for the design and statistical power estimation of future RCTs, potentially helping to overcome gender disparities by explicitly addressing sex differences and enabling studies with equal representation of men and women. Finally, it provides a foundation for follow-up studies into the role of different risk factors, making it a valuable tool to analyze personalized holistic AF management without any costs. We have made the model and source code freely available on https://github.com/HeijmanLab/ to allow others to use and expand this novel approach.

## Conclusions

We developed a novel patient-level AF model that captures the individual dynamic AF development and progression during the entire lifetime of a virtual patient. The model reproduces many important clinical characteristics and population-level metrics of AF in virtual cohorts. V-RCTs simulated by the model revealed how different AF screening strategies can result in heterogenous AF detection rates and benefits for stroke reduction, highlighting potential benefits of AF screening combined with anticoagulation therapy in populations suspected of delayed AF diagnosis. The novel patient-level AF model is expected to enable future studies in the field of basic mechanisms, risk stratification and comprehensive management of AF.

## Data Availability

The code required to generate all data is made publicly available on an open repository (GitHub) at the time of publication. The link to the code is provided in the manuscript.

## Acknowledgements

The authors thank Adele Benzoni, Eva Schuijt, Fateme Farahani, and Paul Volders for helpful discussions.

## Supplemental material

Supplemental methods and steps of sensitivity analysis

Table S1. Values and Sigma of parameters used in the current model.

Table S2. Initial values of different states in the model.

Table S3. Values and Sigma of parameters changed for LOOP study calibration.

Table S4 Parameters setting for the simulation of different AF screening protocols.

Table S5 The detailed information for study-specific model calibration and model validation

Figure S1. The schematic flowchart showing how the states transition runs in the patient-level AF model.

Figure S2. The workflow for model development, calibration, validation and subsequent application.

Figure S3. Sensitivity analysis showing the impacts of individual model parameters on different model outcomes.

Figure S4. The age-dependent and sex-specific prevalence and incidence of clinical atrial fibrillation (AF).

Figure S5. Sex-specific atrial fibrillation (AF) episode characteristics.

Figure S6. Atrial fibrillation (AF) recurrence plots during a 4-week follow-up after spontaneous cardioversion.

Figure S7. The age-dependent and sex-specific mortality and percentage of deaths across different age groups.

Figure S8. The age-dependent and sex-specific stroke incidence and percentage of strokes across different age groups.

Figure S9. The percentage of deaths prehospital (simulated as the first 24 hours), within the first month, or within the first year after stroke compared with data from clinical studies.

Figure S10. Comparison of 5-year and 25-year stroke rates in populations with or without atrial fibrillation (AF) screening with 3-hour diagnostic criteria for clinical AF, EFFICACY OF ANTICOAGULATION THERAPY AS 90% LESS STROKE INCIDENCE, and normal baseline stroke rates.

Figure S11. Comparison of 5-year and 25-year stroke rates in populations with or without atrial fibrillation (AF) screening with 3-hour diagnostic criteria for clinical AF, efficacy of anticoagulation therapy as 70% less stroke incidence, and 3-FOLD INCREASED BASELINE STROKE RATES.

Figure S12. Comparison of 5-year and 25-year stroke rates in populations with or without atrial fibrillation (AF) screening with 24-HOUR DIAGNOSTIC CRITERIA FOR CLINICAL AF, efficacy of anticoagulation therapy as 70% less stroke incidence, and normal baseline stroke rates.

## References

1. Roth GA, Mensah GA, Johnson CO, Addolorato G, Ammirati E, Baddour LM, Barengo NC, Beaton AZ, Benjamin EJ, Benziger CP, et al. Global Burden of Cardiovascular Diseases and Risk Factors, 1990-2019: Update From the GBD 2019 Study. J Am Coll Cardiol. 2020;76:2982–3021. doi: 10.1016/j.jacc.2020.11.010

2. Hindricks G, Potpara T, Dagres N, Arbelo E, Bax JJ, Blomstrom-Lundqvist C, Boriani G, Castella M, Dan GA, Dilaveris PE, et al. 2020 ESC Guidelines for the diagnosis and management of atrial fibrillation developed in collaboration with the European Association for Cardio-Thoracic Surgery (EACTS): The Task Force for the diagnosis and management of atrial fibrillation of the European Society of Cardiology (ESC) Developed with the special contribution of the European Heart Rhythm Association (EHRA) of the ESC. Eur Heart J. 2021;42:373–498. doi: 10.1093/eurheartj/ehaa612

3. Ziff OJ, Carter PR, McGowan J, Uppal H, Chandran S, Russell S, Bainey KR, Potluri R. The interplay between atrial fibrillation and heart failure on long-term mortality and length of stay: Insights from the, United Kingdom ACALM registry. Int J Cardiol. 2018;252:117–121. doi: 10.1016/j.ijcard.2017.06.033

4. Ceornodolea AD, Bal R, Severens JL. Epidemiology and Management of Atrial Fibrillation and Stroke: Review of Data from Four European Countries. Stroke Res Treat. 2017;2017:8593207. doi: 10.1155/2017/8593207

5. Magnussen C, Niiranen TJ, Ojeda FM, Gianfagna F, Blankenberg S, Njolstad I, Vartiainen E, Sans S, Pasterkamp G, Hughes M, et al. Sex Differences and Similarities in Atrial Fibrillation Epidemiology, Risk Factors, and Mortality in Community Cohorts: Results From the BiomarCaRE Consortium (Biomarker for Cardiovascular Risk Assessment in Europe). Circulation. 2017;136:1588–1597. doi: 10.1161/CIRCULATIONAHA.117.028981

6. Kirchhof P, Camm AJ, Goette A, Brandes A, Eckardt L, Elvan A, Fetsch T, van Gelder IC, Haase D, Haegeli LM, et al. Early Rhythm-Control Therapy in Patients with Atrial Fibrillation. N Engl J Med. 2020;383:1305–1316. doi: 10.1056/NEJMoa2019422

7. Hermans ANL, Pluymaekers N, Lankveld TAR, van Mourik MJW, Zeemering S, Dinh T, den Uijl DW, Luermans J, Vernooy K, Crijns H, et al. Clinical utility of rhythm control by electrical cardioversion to assess the association between self-reported symptoms and rhythm status in patients with persistent atrial fibrillation. Int J Cardiol Heart Vasc. 2021;36:100870. doi: 10.1016/j.ijcha.2021.100870

8. Wineinger NE, Barrett PM, Zhang Y, Irfanullah I, Muse ED, Steinhubl SR, Topol EJ. Identification of paroxysmal atrial fibrillation subtypes in over 13,000 individuals. Heart Rhythm. 2019;16:26–30. doi: 10.1016/j.hrthm.2018.08.012

9. Svendsen JH, Diederichsen SZ, Hojberg S, Krieger DW, Graff C, Kronborg C, Olesen MS, Nielsen JB, Holst AG, Brandes A, et al. Implantable loop recorder detection of atrial fibrillation to prevent stroke (The LOOP Study): a randomised controlled trial. Lancet. 2021;398:1507–1516. doi: 10.1016/S0140-6736(21)01698-6

10. Svennberg E, Friberg L, Frykman V, Al-Khalili F, Engdahl J, Rosenqvist M. Clinical outcomes in systematic screening for atrial fibrillation (STROKESTOP): a multicentre, parallel group, unmasked, randomised controlled trial. Lancet. 2021;398:1498–1506. doi: 10.1016/S0140-6736(21)01637-8

11. Healey JS, Lopes RD, Granger CB, Alings M, Rivard L, McIntyre WF, Atar D, Birnie DH, Boriani G, Camm AJ, et al. Apixaban for Stroke Prevention in Subclinical Atrial Fibrillation. N Engl J Med. 2023. doi: 10.1056/NEJMoa2310234

12. Kirchhof P, Toennis T, Goette A, Camm AJ, Diener HC, Becher N, Bertaglia E, Blomstrom Lundqvist C, Borlich M, Brandes A, et al. Anticoagulation with Edoxaban in Patients with Atrial High-Rate Episodes. N Engl J Med. 2023;389:1167–1179. doi: 10.1056/NEJMoa2303062

13. McIntyre WF, Benz AP, Becher N, Healey JS, Granger CB, Rivard L, Camm AJ, Goette A, Zapf A, Alings M, et al. Direct Oral Anticoagulants for Stroke Prevention in Patients with Device-Detected Atrial Fibrillation: A Study-Level Meta-Analysis of the NOAH-AFNET 6 and ARTESiA Trials. Circulation. 2023. doi: 10.1161/CIRCULATIONAHA.123.067512

14. Heijman J, Sutanto H, Crijns H, Nattel S, Trayanova NA. Computational models of atrial fibrillation: achievements, challenges, and perspectives for improving clinical care. Cardiovasc Res. 2021;117:1682–1699. doi: 10.1093/cvr/cvab138

15. Trayanova NA, Lyon A, Shade J, Heijman J. Computational modeling of cardiac electrophysiology and arrhythmogenesis: toward clinical translation. Physiol Rev. 2024;104:1265–1333. doi: 10.1152/physrev.00017.2023

16. Lyth J, Svennberg E, Bernfort L, Aronsson M, Frykman V, Al-Khalili F, Friberg L, Rosenqvist M, Engdahl J, Levin LA. Cost-effectiveness of population screening for atrial fibrillation: the STROKESTOP study. Eur Heart J. 2023;44:196–204. doi: 10.1093/eurheartj/ehac547

17. Aronsson M, Svennberg E, Rosenqvist M, Engdahl J, Al-Khalili F, Friberg L, Frykman-Kull V, Levin LA. Cost-effectiveness of mass screening for untreated atrial fibrillation using intermittent ECG recording. Europace. 2015;17:1023–1029. doi: 10.1093/europace/euv083

18. Van Gelder IC, Healey JS, Crijns H, Wang J, Hohnloser SH, Gold MR, Capucci A, Lau CP, Morillo CA, Hobbelt AH, et al. Duration of device-detected subclinical atrial fibrillation and occurrence of stroke in ASSERT. Eur Heart J. 2017;38:1339–1344. doi: 10.1093/eurheartj/ehx042

19. De With RR, Erkuner O, Rienstra M, Nguyen BO, Korver FWJ, Linz D, Cate Ten H, Spronk H, Kroon AA, Maass AH, et al. Temporal patterns and short-term progression of paroxysmal atrial fibrillation: data from RACE V. Europace. 2020;22:1162–1172. doi: 10.1093/europace/euaa123

20. Nguyen BO, Weberndorfer V, Crijns HJ, Geelhoed B, Ten Cate H, Spronk H, Kroon A, De With R, Al-Jazairi M, Maass AH, et al. Prevalence and determinants of atrial fibrillation progression in paroxysmal atrial fibrillation. Heart. 2022;109:186–194. doi: 10.1136/heartjnl-2022-321027

21. Pluymaekers N, Dudink E, Luermans J, Meeder JG, Lenderink T, Widdershoven J, Bucx JJJ, Rienstra M, Kamp O, Van Opstal JM, et al. Early or Delayed Cardioversion in Recent-Onset Atrial Fibrillation. N Engl J Med. 2019;380:1499–1508. doi: 10.1056/NEJMoa1900353

22. Hart RG, Pearce LA, Aguilar MI. Meta-analysis: antithrombotic therapy to prevent stroke in patients who have nonvalvular atrial fibrillation. Ann Intern Med. 2007;146:857–867. doi: 10.7326/0003-4819-146-12-200706190-00007

23. Ruff CT, Giugliano RP, Braunwald E, Hoffman EB, Deenadayalu N, Ezekowitz MD, Camm AJ, Weitz JI, Lewis BS, Parkhomenko A, et al. Comparison of the efficacy and safety of new oral anticoagulants with warfarin in patients with atrial fibrillation: a meta-analysis of randomised trials. Lancet. 2014;383:955–962. doi: 10.1016/S0140-6736(13)62343-0

24. Bonhorst D, Mendes M, Adragao P, De Sousa J, Primo J, Leiria E, Rocha P. Prevalence of atrial fibrillation in the Portuguese population aged 40 and over: the FAMA study. Rev Port Cardiol. 2010;29:331–350.

25. Lake FR, Cullen KJ, de Klerk NH, McCall MG, Rosman DL. Atrial fibrillation and mortality in an elderly population. Aust N Z J Med. 1989;19:321–326. doi: 10.1111/j.1445-5994.1989.tb00271.x

26. Wolf PA, Abbott RD, Kannel WB. Atrial fibrillation as an independent risk factor for stroke: the Framingham Study. Stroke. 1991;22:983–988. doi: 10.1161/01.str.22.8.983

27. Naccarelli GV, Varker H, Lin J, Schulman KL. Increasing prevalence of atrial fibrillation and flutter in the United States. Am J Cardiol. 2009;104:1534–1539. doi: 10.1016/j.amjcard.2009.07.022

28. Go AS, Hylek EM, Phillips KA, Chang Y, Henault LE, Selby JV, Singer DE. Prevalence of diagnosed atrial fibrillation in adults: national implications for rhythm management and stroke prevention: the AnTicoagulation and Risk Factors in Atrial Fibrillation (ATRIA) Study. JAMA. 2001;285:2370–2375. doi: 10.1001/jama.285.18.2370

29. Miyasaka Y, Barnes ME, Gersh BJ, Cha SS, Bailey KR, Abhayaratna WP, Seward JB, Tsang TS. Secular trends in incidence of atrial fibrillation in Olmsted County, Minnesota, 1980 to 2000, and implications on the projections for future prevalence. Circulation. 2006;114:119–125. doi: 10.1161/CIRCULATIONAHA.105.595140

30. Rietbrock S, Heeley E, Plumb J, van Staa T. Chronic atrial fibrillation: Incidence, prevalence, and prediction of stroke using the Congestive heart failure, Hypertension, Age >75, Diabetes mellitus, and prior Stroke or transient ischemic attack (CHADS2) risk stratification scheme. Am Heart J. 2008;156:57–64. doi: 10.1016/j.ahj.2008.03.010

31. Heeringa J, van der Kuip DA, Hofman A, Kors JA, van Herpen G, Stricker BH, Stijnen T, Lip GY, Witteman JC. Prevalence, incidence and lifetime risk of atrial fibrillation: the Rotterdam study. Eur Heart J. 2006;27:949–953. doi: 10.1093/eurheartj/ehi825

32. Majeed A, Moser K, Carroll K. Trends in the prevalence and management of atrial fibrillation in general practice in England and Wales, 1994-1998: analysis of data from the general practice research database. Heart. 2001;86:284–288. doi: 10.1136/heart.86.3.284

33. Murphy NF, Simpson CR, Jhund PS, Stewart S, Kirkpatrick M, Chalmers J, MacIntyre K, McMurray JJ. A national survey of the prevalence, incidence, primary care burden and treatment of atrial fibrillation in Scotland. Heart. 2007;93:606–612. doi: 10.1136/hrt.2006.107573

34. Piccini JP, Hammill BG, Sinner MF, Jensen PN, Hernandez AF, Heckbert SR, Benjamin EJ, Curtis LH. Incidence and prevalence of atrial fibrillation and associated mortality among Medicare beneficiaries, 1993-2007. Circ Cardiovasc Qual Outcomes. 2012;5:85–93. doi: 10.1161/CIRCOUTCOMES.111.962688

35. Chugh SS, Havmoeller R, Narayanan K, Singh D, Rienstra M, Benjamin EJ, Gillum RF, Kim YH, McAnulty JH, Jr., Zheng ZJ, et al. Worldwide epidemiology of atrial fibrillation: a Global Burden of Disease 2010 Study. Circulation. 2014;129:837–847. doi: 10.1161/CIRCULATIONAHA.113.005119

36. Wilke T, Groth A, Mueller S, Pfannkuche M, Verheyen F, Linder R, Maywald U, Bauersachs R, Breithardt G. Incidence and prevalence of atrial fibrillation: an analysis based on 8.3 million patients. Europace. 2013;15:486–493. doi: 10.1093/europace/eus333

37. Dong XJ, Wang BB, Hou FF, Jiao Y, Li HW, Lv SP, Li FH. Global burden of atrial fibrillation/atrial flutter and its attributable risk factors from 1990 to 2019. Europace. 2023;25:793–803. doi: 10.1093/europace/euac237

38. Alonso A, Agarwal SK, Soliman EZ, Ambrose M, Chamberlain AM, Prineas RJ, Folsom AR. Incidence of atrial fibrillation in whites and African-Americans: the Atherosclerosis Risk in Communities (ARIC) study. Am Heart J. 2009;158:111–117. doi: 10.1016/j.ahj.2009.05.010

39. Gibbs H, Freedman B, Rosenqvist M, Virdone S, Mahmeed WA, Ambrosio G, Camm AJ, Jacobson B, Jerjes-Sanchez C, Kayani G, et al. Clinical Outcomes in Asymptomatic and Symptomatic Atrial Fibrillation Presentations in GARFIELD-AF: Implications for AF Screening. Am J Med. 2021;134:893–901 e811. doi: 10.1016/j.amjmed.2021.01.017

40. Nieuwlaat R, Capucci A, Camm AJ, Olsson SB, Andresen D, Davies DW, Cobbe S, Breithardt G, Le Heuzey JY, Prins MH, et al. Atrial fibrillation management: a prospective survey in ESC member countries: the Euro Heart Survey on Atrial Fibrillation. Eur Heart J. 2005;26:2422–2434. doi: 10.1093/eurheartj/ehi505

41. Verma A, Champagne J, Sapp J, Essebag V, Novak P, Skanes A, Morillo CA, Khaykin Y, Birnie D. Discerning the incidence of symptomatic and asymptomatic episodes of atrial fibrillation before and after catheter ablation (DISCERN AF): a prospective, multicenter study. JAMA Intern Med. 2013;173:149–156. doi: 10.1001/jamainternmed.2013.1561

42. Heijman J, Luermans J, Linz D, van Gelder IC, Crijns H. Risk Factors for Atrial Fibrillation Progression. Card Electrophysiol Clin. 2021;13:201–209. doi: 10.1016/j.ccep.2020.10.011

43. Pritchett EL, DaTorre SD, Platt ML, McCarville SE, Hougham AJ. Flecainide acetate treatment of paroxysmal supraventricular tachycardia and paroxysmal atrial fibrillation: dose-response studies. The Flecainide Supraventricular Tachycardia Study Group. J Am Coll Cardiol. 1991;17:297–303. doi: 10.1016/s0735-1097(10)80090-7

44. Atarashi H, Ogawa S, Inoue H, Hamada C, Flecainide Atrial Fibrillation I. Dose-response effect of flecainide in patients with symptomatic paroxysmal atrial fibrillation and/or flutter monitored with trans-telephonic electrocardiography: a multicenter, placebo-controlled, double-blind trial. Circ J. 2007;71:294–300. doi: 10.1253/circj.71.294

45. van der Velden RMJ, Pluymaekers N, Dudink E, Luermans J, Meeder JG, Heesen WF, Lenderink T, Widdershoven J, Bucx JJJ, Rienstra M, et al. Mobile health adherence for the detection of recurrent recent-onset atrial fibrillation. Heart. 2022;109:26–33. doi: 10.1136/heartjnl-2022-321346

46. Modig K, Talback M, Ziegler L, Ahlbom A. Temporal trends in incidence, recurrence and prevalence of stroke in an era of ageing populations, a longitudinal study of the total Swedish population. BMC Geriatr. 2019;19:31. doi: 10.1186/s12877-019-1050-1

47. Life tables. In: World Health Organization; 2019.

48. Deaths by main cause of death. In: Statistics Netherlands(CBS); 2019.

49. Vaartjes I, O’Flaherty M, Capewell S, Kappelle J, Bots M. Remarkable decline in ischemic stroke mortality is not matched by changes in incidence. Stroke. 2013;44:591–597. doi: 10.1161/STROKEAHA.112.677724

50. Vaartjes I, Reitsma JB, de Bruin A, Berger-van Sijl M, Bos MJ, Breteler MM, Grobbee DE, Bots ML. Nationwide incidence of first stroke and TIA in the Netherlands. Eur J Neurol. 2008;15:1315–1323. doi: 10.1111/j.1468-1331.2008.02309.x

51. Andersson T, Magnuson A, Bryngelsson IL, Frobert O, Henriksson KM, Edvardsson N, Poci D. All-cause mortality in 272,186 patients hospitalized with incident atrial fibrillation 1995-2008: a Swedish nationwide long-term case-control study. Eur Heart J. 2013;34:1061–1067. doi: 10.1093/eurheartj/ehs469

52. Healey JS, Connolly SJ, Gold MR, Israel CW, Van Gelder IC, Capucci A, Lau CP, Fain E, Yang S, Bailleul C, et al. Subclinical atrial fibrillation and the risk of stroke. N Engl J Med. 2012;366:120–129. doi: 10.1056/NEJMoa1105575

53. Chang ET, Lin YT, Galla T, Clayton RH, Eatock J. A Stochastic Individual-Based Model of the Progression of Atrial Fibrillation in Individuals and Populations. PLoS One. 2016;11:e0152349. doi: 10.1371/journal.pone.0152349

54. Khurshid S, Chen W, Singer DE, Atlas SJ, Ashburner JM, Choi JG, Hur C, Ellinor PT, McManus DD, Chhatwal J, et al. Comparative Clinical Effectiveness of Population-Based Atrial Fibrillation Screening Using Contemporary Modalities: A Decision-Analytic Model. J Am Heart Assoc. 2021;10:e020330. doi: 10.1161/JAHA.120.020330

55. Diederichsen SZ, Haugan KJ, Kronborg C, Graff C, Hojberg S, Kober L, Krieger D, Holst AG, Nielsen JB, Brandes A, et al. Comprehensive Evaluation of Rhythm Monitoring Strategies in Screening for Atrial Fibrillation: Insights From Patients at Risk Monitored Long Term With an Implantable Loop Recorder. Circulation. 2020;141:1510–1522. doi: 10.1161/CIRCULATIONAHA.119.044407

56. Lubitz SA, Atlas SJ, Ashburner JM, Lipsanopoulos ATT, Borowsky LH, Guan W, Khurshid S, Ellinor PT, Chang Y, McManus DD, et al. Screening for Atrial Fibrillation in Older Adults at Primary Care Visits: VITAL-AF Randomized Controlled Trial. Circulation. 2022;145:946–954. doi: 10.1161/CIRCULATIONAHA.121.057014

57. Gruwez H, Verbrugge FH, Proesmans T, Evens S, Vanacker P, Rutgers MP, Vanhooren G, Bertrand P, Pison L, Haemers P, et al. Smartphone-based atrial fibrillation screening in the general population: feasibility and impact on medical treatment. Eur Heart J Digit Health. 2023;4:464–472. doi: 10.1093/ehjdh/ztad054

